# Real-world effectiveness of Ad26.COV2.S adenoviral vector vaccine for COVID-19

**DOI:** 10.1101/2021.04.27.21256193

**Authors:** Juan Corchado-Garcia, David Puyraimond-Zemmour, Travis Hughes, Tudor Cristea-Platon, Patrick Lenehan, Colin Pawlowski, Sairam Bade, John C. O’Horo, Gregory J. Gores, Amy W. Williams, Andrew D. Badley, John Halamka, Abinash Virk, Melanie D. Swift, Tyler Wagner, Venky Soundararajan

## Abstract

In light of the massive and rapid vaccination campaign against COVID-19, continuous real-world effectiveness and safety assessment of the FDA-authorized vaccines is critical to amplify transparency, build public trust, and ultimately improve overall health outcomes. In this study, we leveraged large-scale longitudinal curation of electronic health records (EHRs) from the multi-state Mayo Clinic health system (MN, AZ, FL, WN, IA). We compared the infection rate of 2,195 individuals who received a single dose of the Ad26.COV2.S vaccine from Johnson & Johnson (J&J) to the infection rate of 21,950 unvaccinated, propensity-matched individuals between February 27th and April 14th 2021. Of the 1,779 vaccinated individuals with at least two weeks of follow-up, only 3 (0.17%) tested positive for SARS-CoV-2 15 days or more after vaccination compared to 128 of 17,744 (0.72%) unvaccinated individuals (4.34 fold reduction rate). This corresponds to a vaccine effectiveness of 76.7% (95% CI: 30.3-95.3%) in preventing SARS-CoV-2 infection with onset at least two weeks after vaccination. This data is consistent with the clinical trial-reported efficacy of Ad26.COV2.S in preventing moderate to severe COVID-19 with onset at least 14 days after vaccine administration (66.9%; 95% CI: 59.0-73.4%). Due to the recent authorization of the Ad26.COV2.S vaccine, there are not yet enough hospitalizations, ICU admissions, or deaths within this cohort to robustly assess the effect of vaccination on COVID-19 severity, but these outcomes will be continually assessed in near-real-time with our platform. Collectively, this study provides further evidence that a single dose of Ad26.COV2.S is highly effective in preventing SARS-CoV-2 infection and reaffirms the urgent need to continue mass vaccination efforts globally.

## Introduction

To date, there have been over 145 million cases of COVID-19 worldwide with over 3 million associated deaths ^1^. Following the Emergency Use Authorizations by the Food and Drug Administration (FDA) on February 27, 2021, more than 6.8 million doses of the Ad26.COV2.S COVID-19 vaccine (Johnson & Johnson, Janssen) have been administered in the United States^2^. This vaccine consists of a single dose injection of a recombinant, replication-incompetent human adenovirus type 26 vector encoding the SARS-CoV-2 spike (S) protein. A recent phase 3 trial has demonstrated its effectiveness (66.9%, 95% confidence interval [CI], 59.0 to 73.4) and safety profile ^3^. Self-resolving mild to moderate adverse effects were common in vaccinated participants, and serious adverse effects occurred rarely, with a frequency comparable to placebo^3^.

As the vaccines continue to be administered more broadly, it is critical to continuously evaluate safety and effectiveness data for various reasons. For example, the interpretation of vaccine trial outcomes is inherently limited by how representative the studied population is of the broader population which will ultimately receive the vaccine. Further, effectiveness is a dynamic process following which can be impacted by viral evolution. Variants with mutations in the S protein regularly arise and may have the potential to escape the immune response triggered by the vaccine. Finally, the fraction of the population which has been vaccinated can influence the observed effectiveness through herd immunity.

Continuous safety assessment is equally, if not more important. Treating a previously healthy population requires extremely strong evidence of safety (“do not harm” principle), as illustrated by the recent brief pause of Ad26.COV2.S use for safety review following reports of very rare thrombotic thrombocytopenia after vaccination ^2^. While systems such as the Vaccine Adverse Event Reporting System (VAERS) are quite valuable in unearthing rare or ultra rare side effects at the incipient stages ^4^, they rely on active reporting, inherently biased and without appropriate control populations or any medical history on the associated patients.

We have previously leveraged recent advances in deep neural networks to perform high throughput machine-augmented curation of electronic health record (EHR) systems ^5,6^. This has enabled the rapid assessment of real world effectiveness and safety of the mRNA vaccines mRNA-1273 (Moderna) and BNT162b2 (Pfizer/BioNTech), as well as a targeted investigation of cerebral venous sinus thrombosis (CVST) incidence among patients receiving COVID-19 vaccines (including Ad26.COV2.S) within the Mayo Clinic Health System ^7–9^. We believe such approaches are essential for vaccine development, deployment, and most importantly to build trust and transparency for the public.

Here, we expand on this effort to conduct a preliminary assessment of the real-world effectiveness of Ad26.COV2.S vaccination within the multi-state Mayo Clinic Health System (Minnesota, Arizona, Florida, Wisconsin, Iowa) between February 27th and April 14th 2021.

## Results

### Prevention of SARS-CoV-2 infection in a Ad26.COV2.S vaccinated population

Between February 27th and April 14th 2021, a total of 2,195 individuals who met the study inclusion criteria received the Ad26.COV2.s COVID-19 vaccine across the Mayo Clinic network (**Methods** and **Figure 1A**). To evaluate the effectiveness of this vaccine in preventing SARS-CoV-2 infection, we identified a cohort of 21,950 unvaccinated individuals using 1:10 propensity score matching (see **Methods** and **Table 1**). During this time interval, 13 of 2,195 (0.59%) vaccinated individuals have had a positive SARS-CoV-2 PCR test compared to 262 of 21,950 (1.19%) unvaccinated individuals (**Table 2**). The incidence rates of positive SARS-CoV-2 tests in the vaccinated and unvaccinated cohorts were 0.18 and 0.36 cases per 1000 person-days, respectively, indicating an overall vaccine effectiveness of 50.6% (95% CI: 14.0-74.0%) (**Table 2**).

**Table 1.**
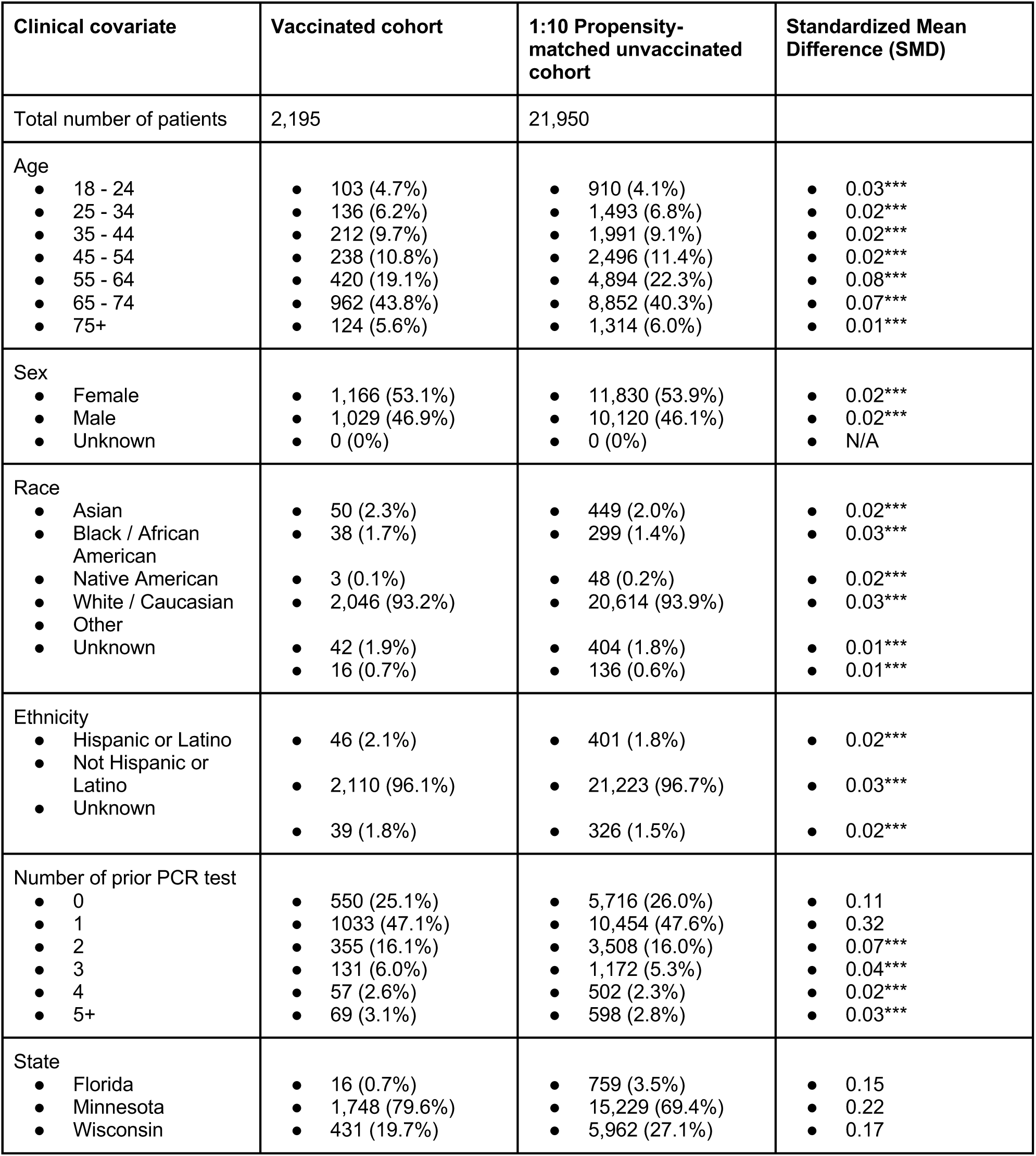
Clinical characteristics of vaccinated and 1:10 propensity-matched unvaccinated cohorts. Covariates for balancing include: (1) Demographics (age, sex, race, ethnicity), (2) Number of prior PCR tests (number of PCR tests that the individual received before December 1, 2020), and (3) Location (zip code). Note that the zip code is matched exactly between the two cohorts, so the proportion of individuals in each state is identical. Highly balanced covariates with Standardized Mean Difference (SMD) < 0.1 are indicated with ***.

| Clinical covariate | Vaccinated cohort | 1:10 Propensity-matched unvaccinated cohort | Standardized Mean Difference (SMD) |
| --- | --- | --- | --- |
| Total number of patients | 2,195 | 21,950 |  |
| Age<br>18 - 24<br>25 - 34<br>35 - 44<br>45 - 54<br>55 - 64<br>65 - 74<br>75+ | 103 (4.7%)<br>136 (6.2%)<br>212 (9.7%)<br>238 (10.8%)<br>420 (19.1%)<br>962 (43.8%)<br>124 (5.6%) | 910 (4.1%)<br>1,493 (6.8%)<br>1,991 (9.1%)<br>2,496 (11.4%)<br>4,894 (22.3%)<br>8,852 (40.3%)<br>1,314 (6.0%) | 0.03***<br>0.02***<br>0.02***<br>0.02***<br>0.08***<br>0.07***<br>0.01*** |
| Sex<br>Female<br>Male<br>Unknown | 1,166 (53.1%)<br>1,029 (46.9%)<br>0 (0%) | 11,830 (53.9%)<br>10,120 (46.1%)<br>0 (0%) | 0.02***<br>0.02***<br>N/A |
| Race<br>Asian<br>Black / African American<br>Native American<br>White / Caucasian<br>Other<br>Unknown | 50 (2.3%)<br>38 (1.7%)<br>3 (0.1%)<br>2,046 (93.2%)<br>42 (1.9%)<br>16 (0.7%) | 449 (2.0%)<br>299 (1.4%)<br>48 (0.2%)<br>20,614 (93.9%)<br>404 (1.8%)<br>136 (0.6%) | 0.02***<br>0.03***<br>0.02***<br>0.03***<br>0.01***<br>0.01*** |
| Ethnicity<br>Hispanic or Latino<br>Not Hispanic or Latino<br>Unknown | 46 (2.1%)<br>2,110 (96.1%)<br>39 (1.8%) | 401 (1.8%)<br>21,223 (96.7%)<br>326 (1.5%) | 0.02***<br>0.03***<br>0.02*** |
| Number of prior PCR test<br>0<br>1<br>2<br>3<br>4<br>5+ | 550 (25.1%)<br>1033 (47.1%)<br>355 (16.1%)<br>131 (6.0%)<br>57 (2.6%)<br>69 (3.1%) | 5,716 (26.0%)<br>10,454 (47.6%)<br>3,508 (16.0%)<br>1,172 (5.3%)<br>502 (2.3%)<br>598 (2.8%) | 0.11<br>0.32<br>0.07***<br>0.04***<br>0.02***<br>0.03*** |
| State<br>Florida<br>Minnesota<br>Wisconsin | 16 (0.7%)<br>1,748 (79.6%)<br>431 (19.7%) | 759 (3.5%)<br>15,229 (69.4%)<br>5,962 (27.1%) | 0.15<br>0.22<br>0.17 |

**Table 2.** SARS-CoV-2 incidence rates in vaccinated and 1:10 propensity-matched unvaccinated cohorts, and corresponding vaccine effectiveness. Incidence is calculated as the number of cases per 1000 person-days. The columns are: **(1) Time Period:** Time period relative to vaccine dose for vaccinated cohort or study enrollment day for unvaccinated cohort; **(2) Vaccinated Incidence Rate:** Number of patients with positive PCR tests in the vaccinated cohort in the time period, divided by the number of at-risk person-days for the vaccinated cohort in the time period; in brackets, the number of cases per 1000 person-days; **(3) Unvaccinated Incidence Rate:** Number of patients with positive PCR tests in the propensity-matched unvaccinated cohort in the time period, divided by the number of at-risk person-days for the propensity-matched unvaccinated cohort in the time period; in brackets, the number of cases per 1000 person-days; **(4) Incidence Rate Ratio:** Vaccinated Incidence Rate divided by Unvaccinated Incidence Rate, along with the exact 95% confidence interval (17), **(5) Vaccine effectiveness:** 100% × (1-Incidence Rate Ratio), along with the 95% confidence interval.

| Time period | Vaccinated cases/person-days [Per 1000 Person-Days] (# Patients Contributing) | Unvaccinated cases/person-days [Per 1000 Person-Days] (# Patients Contributing) | Incidence rate ratio (exact 95% CI) | Vaccine effectiveness (95% CI) |
| --- | --- | --- | --- | --- |
| Day 1 onwards | 13 / 73,809. [0.18] (n = 2,195) | 262 / 735,390. [0.36] (n = 21,950) | 0.49 (0.26, 0.86) | 50.6% (14.0%, 74.0%) |
| Day 8 onwards | 6 / 56,264. [0.11] (n = 2,188) | 173 / 560,124. [0.31] (n = 21,861) | 0.35 (0.12, 0.77) | 65.5% (23.3%, 87.5%) |
| Day 15 onwards | 3 / 41,924. [0.072] (n = 1,779) | 128 / 416,894. [0.31] (n = 17,744) | 0.23 (0.047, 0.7) | 76.7% (30.3%, 95.3%) |

**Figure 1.**
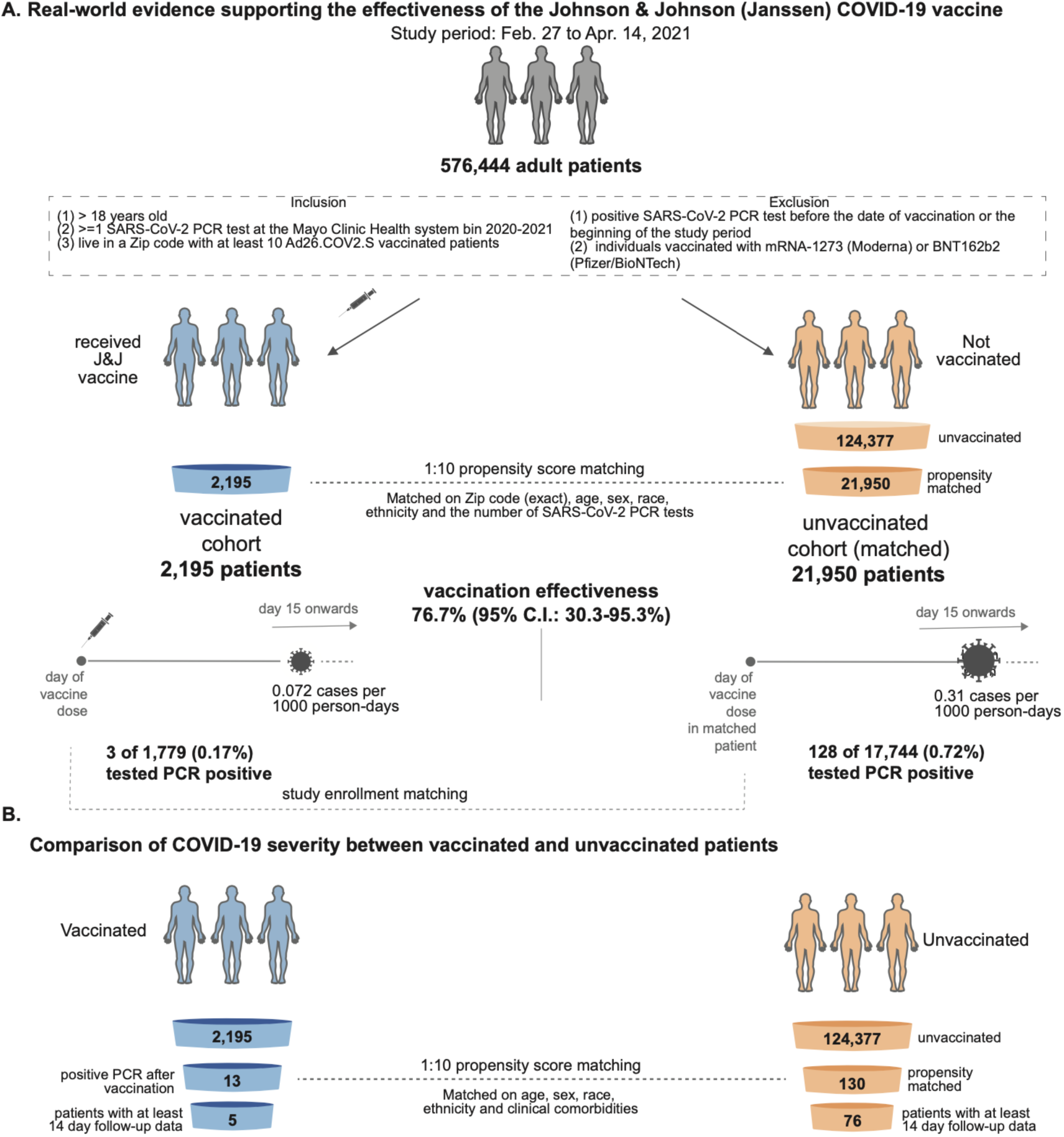
Schematic illustrating the analysis of Ad26.COV2.S (Johnson & Johnson, J&J) vaccine effectiveness by comparing vaccinated and unvaccinated cohorts. **(A)** Analysis of effectiveness in preventing SARS-CoV-2 infection. Out of 126,572 available adult patients during the study period (Feb 27, 2021 to April 14, 2021), 2,195 patients were vaccinated. A 1:10 control cohort of 21,950 patients was designed by propensity-matching for COVID-19 infection risk (e.g. location). Starting 15 days after study enrollment, 3 of 1,779 vaccinated patients tested positive for SARS-COV-2 (by PCR) after vaccination vs 128 of 17,744 unvaccinated individuals, corresponding to an effectiveness of 76.7% achieved two weeks after vaccination. (B) Analysis of effectiveness in preventing severe SARS-CoV-2 infection (hospitalization, ICU admission, mortality). In order to control for age and comorbidities, a 1:10 control cohort of 130 patients was designed by propensity-matching to match the 13 infected individuals that were vaccinated. No difference is observed but too few events were available at the time of publication to provide adequate power for this analysis.

The full effectiveness of Ad26.COV2.S is expected to be achieved after several weeks ^3^. Thus, we next analyzed the incidence rates of positive SARS-CoV-2 tests starting 15 days after the study enrollment date. During this time, 3 of 1,779 (0.17%) vaccinated individuals tested positive compared to 128 of 17,744 (0.72%) unvaccinated individuals. This corresponds to a vaccine effectiveness of 76.7% (95% CI: 30.3-95.3%) in preventing SARS-CoV-2 infection with onset at least two weeks after vaccination (**Table 2**). Consistent with this, Kaplan-Meier analysis to investigate hazard reduction after receiving the Ad26.COV2.S vaccine shows that the curves split approximately 14 days after injection (**Figure 2A-B**).

**Figure 2.**
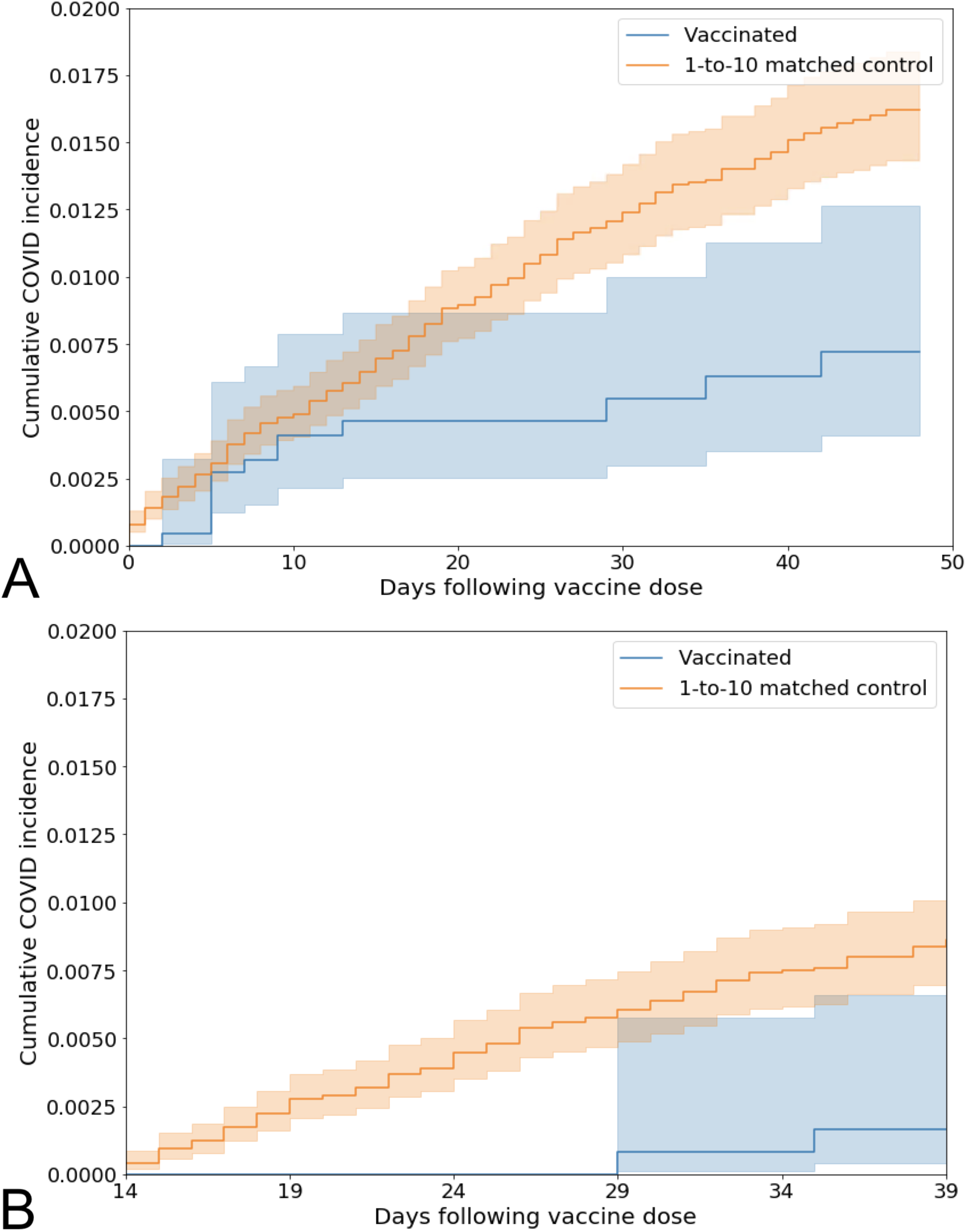
Kaplan Meier analyses to assess cumulative proportional incidence of SARS-CoV-2 infection between vaccinated and unvaccinated individuals. Cumulative proportional incidence at time *t* is the estimated proportion of patients who experience the outcome on or before time *t*, i.e. 1 minus the standard Kaplan-Meier survival estimate. Cumulative proportional incidence of SARS-CoV-2 infection with onset (A) on any day after the date of vaccination (13 / 54,190 vaccinated, 244 / 520,736 unvaccinated; log-rank test p-value 0.0078), or (B) after 14 days from the date of vaccination (3 / 28,847 vaccinated, 120 / 275,908 unvaccinated; log-rank test p-value 0.0035).

Infection can occur after vaccination if an individual is exposed to SARS-CoV-2 shortly after vaccination (i.e., before building immunity) or if the vaccinate fails to elicit protective immunity. In order to distinguish between the two, we analyzed the distribution of time between study enrollment and the fist positive PCR test in these two cohorts **(Figure 3)**. The median time to positive PCR test following vaccination was 7 days, with 10 of 13 infections occurring within the first two weeks. For the unvaccinated individuals, the median time to positive PCR test following enrollment date was 14 days. These results suggest that most infections in the vaccinated cohort are likely due to viral exposure before the recipient was able to develop immunity rather than failure of the vaccine itself.

**Figure 3.**
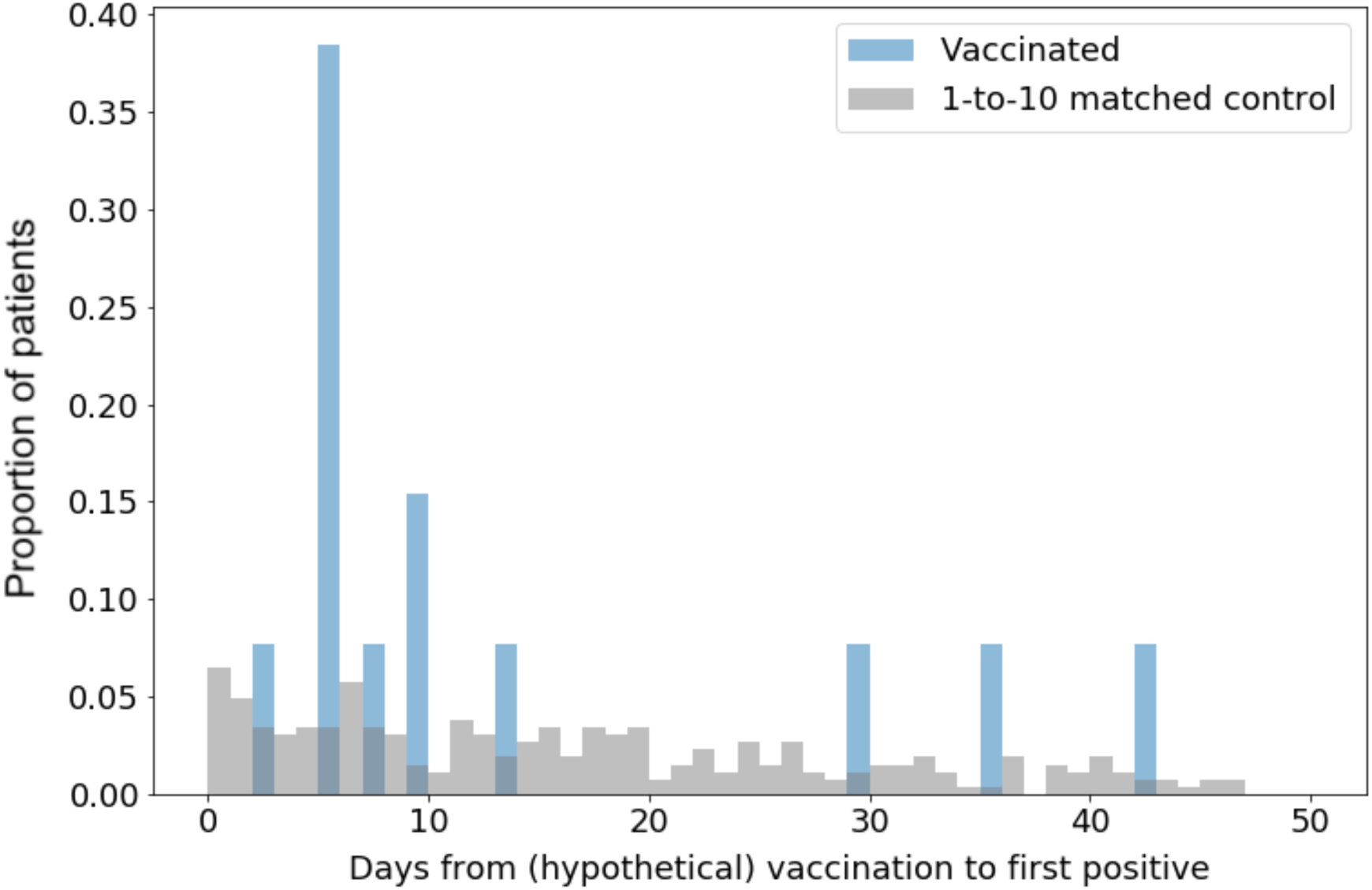
Distribution of the time to the first positive PCR test in Ad26.COV2.S vaccinated individuals (blue) and unvaccinated (grey).

### Rate of severe COVID-19 disease following Ad26.COV2.s vaccination

To understand effectiveness in preventing severe COVID-19 (hospitalization, ICU admission, and death), we first compared the rate of hospitalization, ICU admission and mortality between the 13 infected vaccinated individuals and the 262 infected unvaccinated individuals. No difference in hospitalization rate, ICU admission rates were observed. A difference in mortality trended towards the end of the follow-up period but there was not enough time to reach significance **(Figure 4)**.

**Figure 4.**
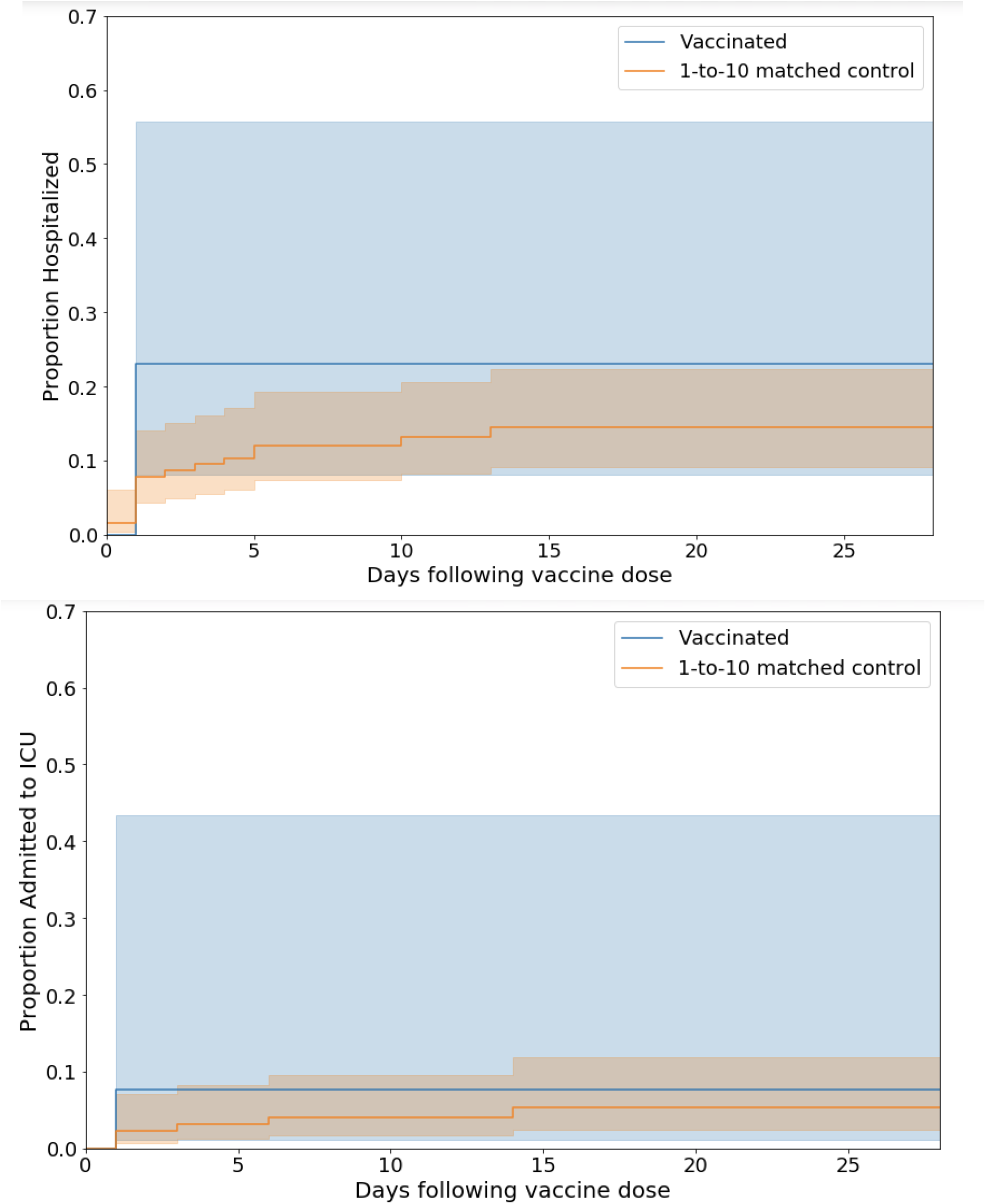
Kaplan Meier analyses to assess COVID-19 disease severity between vaccinated and unvaccinated patients. (A) Hospitalization survival comparison between patients who tested positive for SARS-CoV-2 after being vaccinated (n = 13) versus the 262 patients who tested COVID19 positive and were not vaccinated (log-rank test p-value of 0.31). (B) same as A for ICU admission rate (log-rank test p-value of 0.58).

Currently, vaccination is prioritized towards older and at-risk individuals, which biases the comparison with the unvaccinated cohort (younger, healthier). In order to control for this bias, we designed a cohort of unvaccinated patients matching the 13 vaccinated patents that were infected by SARS-CoV-2 after vaccination **(Figure 1B, Methods**). Using 1:10 propensity score matching, the cohort was matched based on demographics (age, sex, race, ethnicity), and risk factors for severe COVID-19 illness provided by the Centers of Disease Control and Prevention ^10^ **(see Methods)**. We then compared the 14-day rates of hospitalization, ICU admission, and mortality between these cohorts **(Table S3)**. While we did not observe any significant differences in these outcomes, it is important to note that only 5 vaccinated COVID-19 patients had sufficient follow up (14 days) for inclusion in this analysis. This cohort is not yet large enough to robustly assess the impact of Ad26.COV2.S on COVID-19 severity, as we estimate that 149 events within 14 days would be required to detect a difference of 85% in hospitalization, ICU admission, or mortality with a statistical power of 80%.

## Discussion

In this study, we assessed the effectiveness of Ad26.COV2.S COVID-19 vaccine (Johnson & Johnson, Janssen) in a real world setting by analyzing longitudinal health records of 2,195 vaccinated individuals and a 1:10 propensity matched unvaccinated cohort in the multi-state Mayo Clinic Health System (MN, AZ, FL, WN, IA). We observed significant reductions in rates of newly diagnosed COVID-19 infection, with an overall effectiveness of 48.8% (95% CI: 10.8-73.1%) and an effectiveness of 76.1% (95% CI: 28.4-95.1%) starting 14 days after vaccination. These findings are consistent with the results of the phase 3 trial of Ad26.COV2.S, which demonstrated 66.9% efficacy against moderate to severe COVID-19 with onset at least 14 days after vaccination ^3^.

Clinical trials have demonstrated that multiple COVID-19 vaccines, including Ad26.COV2.S, are highly efficacious in reducing the risk of severe illness. However, because only 13 individuals tested positive for SARS-CoV-2 after receiving Ad26.COV2.S in our study, we were underpowered to assess an impact on COVID-19 severity in this population. As the vaccine is administered to more patients, we will continue to assess rates of hospitalization, ICU admission, and mortality among individuals who become infected after Ad26.COV2.S vaccination.

This study has several limitations. First, the study was conducted using data from a single health system, with the vast majority of analyzed individuals residing in Minnesota or Wisconsin. The cohorts, which are over 90% Caucasian and approximately 54% female, are not demographically representative of the broader United States population that is now eligible for vaccination (**Table 1**). Second, the cohort analyzed is relatively small compared to the population analyzed in the phase 3 trial of Ad26.COV2.S and compared to our prior real world analyses of FDA-authorized mRNA vaccines ^3,7^. Finally, we did not assess the safety profile of Ad26.COV2.S here. It should be noted that we have previously performed a targeted investigation which showed that no individuals in the Mayo Clinic health system who received this vaccine have yet developed CVST, but this finding should also be taken in context of the very limited cohort size.

Despite these caveats, this study is the first propensity matched real-world effectiveness assessment of Ad26.COV2.S. Implementation of this framework will allow us to track in real time how the effectiveness of this one-shot vaccine continues to evolve over the coming weeks. The extraction of such data from holistic health records inference is critical in shaping the global race against the ongoing pandemic. This information is particularly important in the context of variant emergence that could potentially escape vaccine-induced immunity. In addition, the recent brief pause in Ad26.COV2.S administration to analyze its safety highlights the value for independent analysis and better communication about vaccines, to ensure transparency and encourage trust in a life-saving procedure for our communities.

## Methods

### Study design and participants

This is a retrospective study of individuals who underwent polymerase chain reaction (PCR) testing for suspected SARS-CoV-2 infection at the Mayo Clinic and hospitals affiliated with the Mayo Clinic Health System. This study was reviewed by the Mayo Clinic Institutional Review Board (IRB) and determined to be exempt from the requirement for IRB approval (45 CFR 46.104d, category 4). Subjects were excluded if they did not have a research authorization on file.

The participant selection algorithm for this study mirrors that outline in our previous real world effectiveness analysis of mRNA COVID-19 vaccines ^7^. Specifically in this retrospective study, patients were included with the following criteria: (1) underwent at least one SARS-CoV-2 polymerase chain reaction (PCR) test at the Mayo Clinic Health system in 2020-2021; (2) at least 18 years old; (3) resides in a local (based on Zip code) in which at least 10 patients who have received the Ad26.COV2.S vaccine.

Exclusion criteria were: (1) positive SARS-CoV-2 PCR test before the date of vaccine administration or the beginning of the study period (Feb 27, 2021) (2) individuals with zero follow-up days after vaccination (i.e. those who received the vaccine dose on the last date of data collection); (3) individuals vaccinated with mRNA-1273 (Moderna) or BNT162b2 (Pfizer/BioNTech) and (4) no research authorization on file.

After applying these inclusion and exclusion criteria, the study population included 2,195 vaccinated and 124,377 unvaccinated patients (**Figure 1A**). Further selection of the unvaccinated cohort was performed to match the vaccinated population: see sections below on the propensity score matching procedure.

### Effectiveness analysis (SARS-CoV-2 infection rate)

#### Propensity score matching procedure defining the unvaccinated cohort

We employed 1:10 propensity score matching ^11^ to construct an unvaccinated cohort similar to the vaccinated cohort with respect to key risk factors for SARS-CoV-2 infection, as was described previously ^7^: (i) geography (zip code of the patient’s residence), (ii) demographics (age, sex, race, ethnicity), (iii) and records of PCR testing (number of negative PCR tests taken before December 1st, 2020. As described previously, the number of negative PCR tests intends to capture a combination of factors including potential exposure levels to COVID-19 along with the willingness and ability to undergo SARS-CoV-2 testing. Propensity scores were obtained by training regularized logistic regression models for each zip code using the software package sklearn v0.20.3 in Python.

Using these propensity scores, we matched each of the 2,195 individuals in the previously defined vaccinated cohort with 10 patients out of the 124,377 eligible unvaccinated individuals, using greedy nearest-neighbor matching without replacement ^12^. It should be noted that if an unvaccinated individual tested positive for SARS-CoV-2 prior to the vaccination date for a potential matched vaccinated individual, this was considered an invalid match; in such cases, the unvaccinated individual was recycled (i.e. made available to potentially match to other vaccinated individuals) and a new unvaccinated individual was selected from the pool.

#### Evaluation of vaccine effectiveness

Vaccine effectiveness was evaluated as described previously ^7^. For each vaccinated individual, the date of study enrollment (Day 0) was defined as the date of vaccine administration. For each unvaccinated individual, the date of study enrollment was defined as the date of the vaccine administration for their matched vaccinated individual.

Cumulative proportional incidence of SARS-CoV-2 infection was compared between vaccinated and unvaccinated patients by Kaplan Meier analysis. Cumulative proportional incidence at time t is the estimated proportion of patients who experience the outcome on or before time t, i.e. 1 minus the standard Kaplan-Meier survival estimate. We considered cumulative incidence starting at Day 1, Day 14 relative to the date of study enrollment (Day 0). Statistical significance was assessed with the log rank test.

We calculated the incidence rate ratio (IRR) of positive SARS-CoV-2 tests between the vaccinated and unvaccinated cohorts. Effectiveness was defined as 100% × (1 - IRR). We considered the incidence rate starting on (i) Day 1 after vaccination, (ii) Day 8 after vaccination, and (iii) Day 15 after vaccination. Incidence rates were defined as the number of patients testing positive for SARS-CoV-2 in the given time period divided by the total number of at-risk person-days contributed in that time period. For each individual, at-risk person-days are defined as the number of days in the time period in which the individual has not yet tested positive for SARS-CoV-2 or died. The IRR was calculated as the incidence rate of the vaccinated cohort divided by the incidence rate of the unvaccinated cohort, and its 95% confidence interval was computed using an exact approach described previously ^13^.

### Severe COVID19 after vaccination

#### Propensity score matching procedure defining the unvaccinated cohort

We applied 1:10 propensity score matching to construct a SARS-CoV-2 positive unvaccinated cohort similar in baseline clinical covariates to the cohort of patients who were vaccinated and subsequently tested positive for SARS-CoV-2. Patients were matched based on demographics (age, sex, race, ethnicity), comorbidities (asthma, cancer, cardiomyopathy, chronic kidney disease, chronic obstructive pulmonary disease, coronary artery disease, heart failure, hypertension, obesity, pregnancy, severe obesity, sickle cell disease, solid organ transplant, stroke / cerebrovascular disease, type 2 diabetes mellitus). This list of comorbidities was derived from the list of risk factors for severe COVID-19 illness provided by the Centers of Disease Control and Prevention ^10^. We used deep neural networks to automatically identify comorbidities from the clinical notes, which are described in the next section. To obtain the propensity scores, we trained a regularized logistic regression model with these features using the software package sklearn v0.20.3 in Python.

Based on these propensity scores, we matched each of the 13 individuals that tested positive for SARS-CoV-2 after vaccination with 130 individuals out of the 262 individuals that tested positive for SARS-CoV-2 and that were not vaccinated, using greedy nearest-neighbor matching without replacement. The resulting cohorts are summarized in **Table S2**, along with the SMDs for the clinical covariates that were balanced upon. There were no significant differences between the two cohorts in any of the clinical covariates that were included in propensity score matching (with SMD < 0.1 for all covariates).

For each SARS-CoV-2 positive patient in both the vaccinated and unvaccinated cohorts, the index date for the analysis (day 0) was taken to be the date of the first positive PCR test. Clinical outcomes at 14 days were compared, including hospital admission, ICU admission, and mortality.

#### Disease severity analysis

Analysis was performed with patients in each cohort with at least 14 days of follow-up after their first positive PCR test (n = 5 vaccinated, 165 unvaccinated). The following parameters were evaluated: (1) 14-day hospital admission rate: Number of patients admitted to the hospital in the two weeks following their positive PCR test, (2) 14-day ICU admission rate: Number of patients admitted to the ICU in the two weeks following their positive PCR test, and (3) 14-day mortality rate: Number of patients deceased in the two weeks following their positive PCR test. For each outcome, we report the relative risk (rate in the vaccinated cohort divided by the rate in the matched unvaccinated cohort), 95% confidence interval for the relative risk, and Fisher’s exact test p-value. Hospital-free and ICU-free survival were also compared via Kaplan-Meier analysis, with statistical significance assessed with the log rank test.

#### Automated clinical data extraction from clinical notes

We used our previously described BERT-based neural network model to identify comorbidities from the electronic health record for each patient and classify the sentiment for the phenotypes that appeared in the clinical notes ^14^. Briefly, we applied a phenotype sentiment classification model that had been trained on 18,500 sentences which achieves an out-of-sample accuracy of 93.6% with precision and recall scores above 95%. This classification model predicts four classes, including: (1) “Yes”: confirmed diagnosis (2) “No”: ruled-out diagnosis, (3) “Maybe”: possibility of disease, and (4) “Other”: alternate context (e.g. family history of disease). For each patient, we applied the sentiment model to the clinical notes in the Mayo Clinic electronic health record. For each comorbidity phenotype, if a patient had at least one mention of the phenotype during the time period with a confidence score of 90% or greater, then the patient was labelled as having the phenotype.

## Data Availability

After publication, the data will be made available upon reasonable requests to the corresponding author. A proposal with detailed description of study objectives and the statistical analysis plan will be needed for evaluation of the reasonability of requests. Deidentified data will be provided after approval from the corresponding author and the Mayo Clinic.

## Author Contributions

VS and TW conceived the study. DZ, JCG, TH and PL wrote the manuscript and reviewed the findings. JCG, DZ, TH, PL, TCP, CP, SB, TW contributed methods, analysis, and software. JCOH, GJG, AWW, ADB, MDS, AV, and JH reviewed the study design, clinical findings, and the manuscript. All authors revised the manuscript.

## Supplementary Material

**Figure S1.**
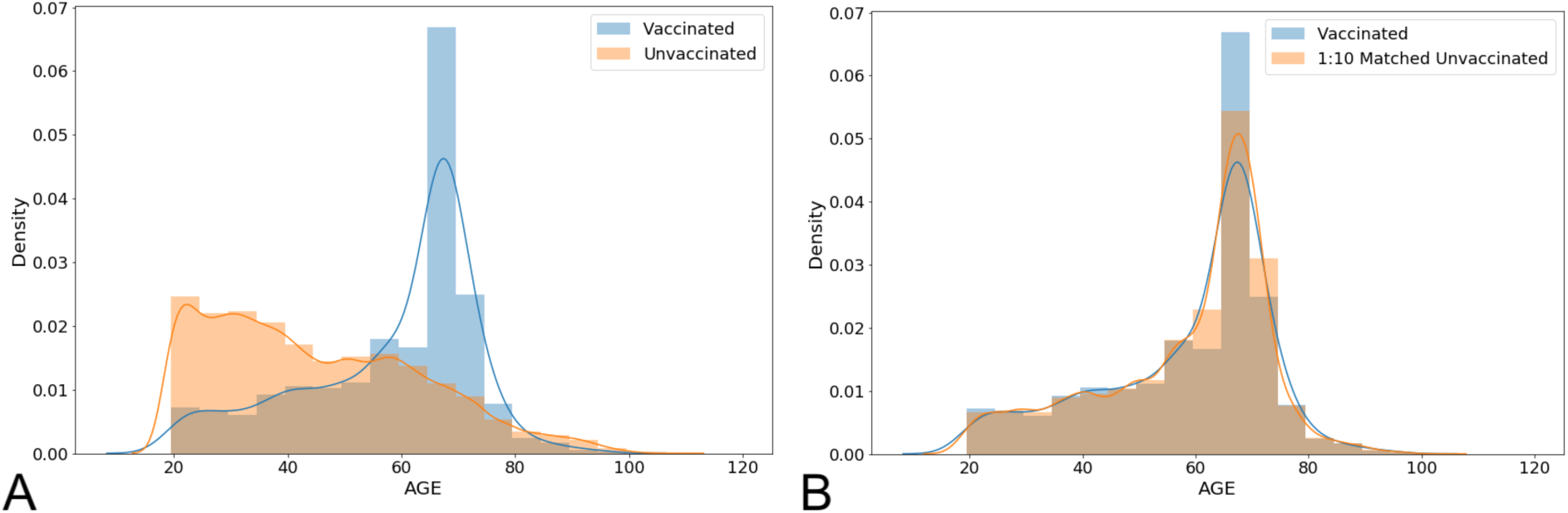
Age distributions for vaccinated and unvaccinated cohorts before and after propensity matching. (A-B) Distribution of ages for vaccinated patients and unvaccinated patients before (A) and after (B) 1:10 matching. These matched cohorts were used to assess vaccine effectiveness in preventing SARS-CoV-2 infection.

**Figure S2.**
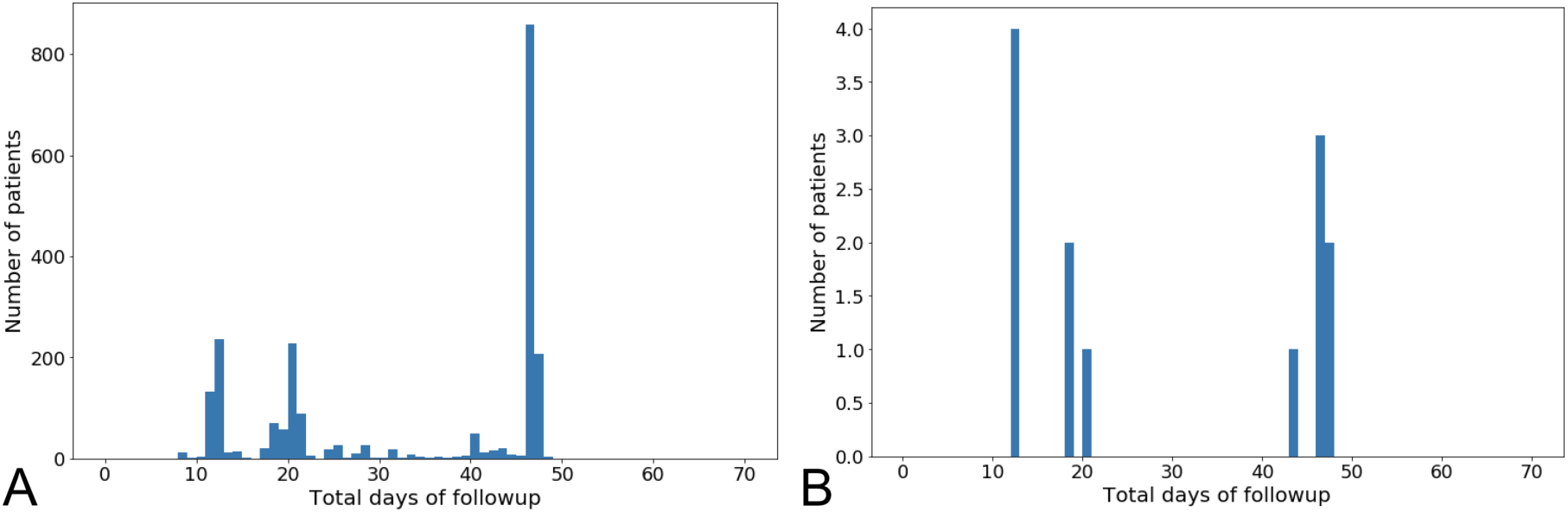
Distributions of follow-up time. (A) Follow-up time (days) for the 2,182 individuals who received a COVID-19 vaccine and did not have a positive SARS-CoV-2 test before their vaccine injection. Total available follow-up time is defined as the number of days from vaccine injection to the final study date (April 14, 2021), including days after COVID-19 diagnosis or death, if applicable. (B) Total available follow-up time (days) for the 13 individuals who received a COVID-19 vaccine subsequently tested positive for SARS-CoV-2 by PCR. Total available follow-up time is defined as the number of days from diagnosis (date of positive SARS-CoV-2 test) to the final study date (April 14, 2021), including days after COVID-19 diagnosis or death, if applicable.

**Table S2.** Clinical characteristics of SARS-CoV-2 positive vaccinated and 1:10 propensity matched unvaccinated cohorts. The SARS-CoV-2 positive vaccinated cohort includes all patients who received the Ad26.COV2.S vaccine and then subsequently received a positive PCR test. The control cohort is a 1:10 propensity-matched cohort derived from the set of unvaccinated patients with a positive PCR test on or after February 27, 2021. Demographics and comorbidities are presented for each cohort, and number of doses is presented for the vaccinated cohort. Comorbidities were determined via neural network models applied to clinical notes for each patient. Highly balanced covariates with Standardized Mean Difference (SMD) < 0.1 are indicated with ***.

| Clinical covariate | SARS-CoV-2 positive vaccinated cohort | 1:10 Propensity-matched SARS-CoV-2 positive unvaccinated cohort | Standardized Mean Difference (SMD) |
| --- | --- | --- | --- |
| Total number of patients | 13 | 130 |  |
| Age<br>18 - 44 years old<br>45 - 64 years old<br>>= 65 years old | 2 (15.4%)<br>5 (38.5%)<br>6 (46.2%) | 14 (10.8%)<br>45 (34.6%)<br>71 (54.6%) | 0.15*<br>0.08***<br>0.17* |
| Sex<br>Female<br>Male<br>Unknown | 5 (38.5%)<br>8 (61.5%)<br>0 (0%) | 51 (39.2%)<br>79 (60.8%)<br>0 (0%) | 0.02***<br>0.02***<br>N/A |
| Race<br>Asian<br>Black / African American<br>Native American<br>White / Caucasian<br>Other<br>Unknown | Race<br>0 (0%)<br>1 (7.7%)<br>0 (0%)<br>12 (92.3%)<br>0 (0%)<br>0 (0%) | Race<br>0 (0%)<br>1 (0.8%)<br>0 (0%)<br>129 (99.2%)<br>0 (0%)<br>0 (0%) | Race<br>N/A<br>0.59<br>N/A<br>0.59<br>N/A<br>N/A |
| Ethnicity<br>Hispanic or Latino<br>Not Hispanic or Latino<br>Unknown | 0 (0%)<br>13 (100%)<br>Unknown | 2 (1.5%)<br>127 (97.7%)<br>Unknown | N/A<br>0.16*<br>Unknown |
| Comorbidities<br>Asthma<br>Cancer<br>Cardiomyopathy<br>Chronic Kidney Disease<br>Chronic Obstructive Pulmonary Disease<br>Coronary Artery Disease<br>Heart failure<br>Hypertension<br>Obesity<br>Pregnancy<br>Severe Obesity<br>Sickle Cell Disease<br>Solid Organ Transplant<br>Stroke / Cerebrovascular Disease<br>Type 2 Diabetes Mellitus | 3 (23.1%)<br>1 (7.7%)<br>0 (0%)<br>2 (15.4%)<br>2 (15.4%)<br>3 (23.1%)<br>1 (7.7%)<br>7 (53.8%)<br>4 (30.8%)<br>0 (0%)<br>0 (0%)<br>0 (0%)<br>0 (0%)<br>0 (0%)<br>0 (0%) | 5 (3.8%)<br>15 (11.5%)<br>5 (3.8%)<br>14 (10.8%)<br>6 (4.6%)<br>13 (10%)<br>13 (10%)<br>66 (50.8%)<br>37 (28.5%)<br>0 (0%)<br>2 (1.5%)<br>0 (0%)<br>0 (0%)<br>3 (2.3%)<br>10 (7.7%) | 0.86<br>0.12*<br>0.21*<br>0.15*<br>0.47<br>0.41<br>0.08***<br>0.06***<br>0.05***<br>N/A<br>0.13*<br>N/A<br>N/A<br>0.16*<br>0.30 |

**Table S3.** 14-day rates of hospitalization, ICU admission, and mortality for vaccinated vs 1:10 propensity-matched unvaccinated COVID-19 patients. Patients were considered eligible for analysis if they had at least 14 days of follow-up after COVID-19 diagnosis as defined by a positive SARS-CoV-2 PCR test. For each outcome, the relative risk (and its 95% confidence interval) and Fisher exact test p-value are used to compare the rates between vaccinated and unvaccinated patients. To indicate statistical significance, * denotes p-value < 0.05, ** denotes p-value < 0.01.

| **Outcome** | **J&J 1 dose,**<br>**COVID positive**<br>**(13 patients)** | **Matched**<br>**unvaccinated,**<br>**COVID positive**<br>**(130 patients)** | **Relative Risk**<br>**(95% CI)** | **Fisher Exact test**<br>**p-value** |
| --- | --- | --- | --- | --- |
| Number of patients with at least 14 days of follow-up | 5 | 76 |  |  |
| 14-Day Hospital admission rate | 1/5 (20%) | 13/76 (17.1%) | 0.68 (0.12, 2.99) | 0.68 |
| 14-Day ICU admission rate | 0/5 (0%) | 6/76 (7.9%) | 0 (0, 7.09) | 0.59 |
| 14-Day Mortality rate | 0/5 (0%) | 1/76 (1.3%) | 0 (0, 42.76) | 1 |

